# Comparing Human and Large Language Model Responses to Patients’ Online Questions: Towards Multi-dimensional Patient-centered Support

**DOI:** 10.64898/2026.07.15.26355314

**Authors:** Md Alomgeer Hussein, Rajmi Doshi, Lu He, Tera L. Reynolds

## Abstract

Patients and caregivers seek informational and emotional support throughout medical care, especially when interpreting unfamiliar laboratory test results. Although resources such as patient portals and online health communities (OHCs) help address questions, gaps remain. The emergence of large language models (LLMs) offers the potential to be a complementary source of support to assist patients and caregivers in understanding and using their test results. The objective of our study is to empirically compare LLM responses to patients’ online questions containing their laboratory test results to responses written by peers in an OHC. We compared the 519 peer replies to 122 laboratory test-related posts from an OHC to 488 responses generated from four LLMs using mixed computational and qualitative methods. LLMs frequently provided clear explanations of medical terminology and structured interpretations of numeric results but were longer and less readable. Peers offered more personalized, context-specific emotional support. Overall, LLMs have the potential to complement peer responses in OHCs, but require greater emotional depth, reasoning transparency, and alignment with community norms.

## Introduction

Patients have easier and faster access to their medical records than ever before through patient portals. At the same time, patients also assume more responsibility for their health, often making decisions outside of the formal healthcare systems (e.g., deciding when a concern warrants a visit to a doctor)^1,2^. This often leads to uncertainty and, subsequently, help-seeking on the part of the patients. Existing research suggests that patients engage an “ecosystem of support” when seeking help—friends and family, healthcare providers, and online information and communities (e.g., Facebook groups)—often sharing medical information such as laboratory test results in this process^2,3^.

With the major advancements in large language models (LLMs) in the past two years, there is anecdotal evidence of patients adding these tools to their ecosystem of support, particularly when all other avenues have been exhausted^4^. Unlike traditional resources, LLMs can offer interactive, personalized guidance that supports deeper learning. Given these advantages, LLMs are likely to play an increasingly prominent role in patients’ healthcare journeys. The projected wider adoption of LLMs for health information seeking and the unclear benefits and unintended consequences present an urgent need for research investigating the potential of LLMs for this purpose. While recent research examined whether LLMs can provide medically correct and safe responses to patients’ questions^5,6^, reference relevant medical evidence^7,8^, and summarize medical evidence^9^, many open questions remain. For example, while research has shown the importance of patient-facing content being readable^10^, personalized^11^, and emotionally supportive^2,12–14^, less is known about the extent to which LLMs meet these needs. In addition, a larger question remains – how might LLMs fit into the existing ecosystem of support to maximize benefits while minimizing risks?

We addressed these gaps through a multi-method study that utilized natural language processing and qualitative approaches to investigate the types of support provided by LLMs compared to the types of support provided by peer patients using anonymous questions and threaded replies from an online health community (OHC). Our approach shifts from the clinical-centered approach focused on the accuracy of medical advice to a patient-centered approach focused on whether the responses can provide holistic support to patients. A comprehensive understanding of the potential for LLM-based tools on OHCs is especially important since one of the reasons that people turn to OHCs is not having immediate access to traditional healthcare resources^2^, suggesting that tools provided on OHCs could harm already disadvantaged patient populations – those with gaps in their ecosystems of support – if not carefully designed.

## Methods

We evaluated four popular LLMs, GPT-3.5-Turbo-1106, GPT-4o, DeepSeek-R1, and Mistral-7B-Instruct-v0.2 by providing anonymous questions that contain an individual’s laboratory test results that were posted to an OHC and comparing the LLM-generated responses against OHC user responses. We focused on laboratory test results as a starting point because they are often viewed through patient portals without support from medical professionals and are common sources of questions^15–17^. Beyond accurate medical information, patients and their caregivers often look for emotional support, including reassurance and empathy, particularly during challenging treatments^2,11–13^. UMBC’s Institutional Review Board deemed this non-human subject research.

### OHC Dataset

We utilized a previously collected and curated dataset from one OHC^2^. Briefly, the data were collected from a popular OHC that had a membership exceeding one million unique users across more than 400 forums at the time of data collection. Patients and caregivers exchanged informational and emotional support through asking and answering questions and sharing stories and updates. Topics within these forums varied widely, from specific health issues like diabetes and thyroid disorders to wellness topics such as nutrition advice. While OHC users came from many different countries, most were from the U.S. The conversations were primarily between patients and caregivers; however, there was also some participation from healthcare professionals (e.g., nurses) who provided expert insights.

The foundation of our study was a curated dataset of question posts in English that were manually reviewed in a previous study^2^ to confirm that they include the question poster’s laboratory test results and one or more questions. From the 321 posts in that dataset, we retained the 122 with threaded replies (N=519 comments). This dataset was curated from a random sample of 1,000 question posts taken from a dataset containing 60,000 posts with indicators that they contain laboratory test results (e.g., a number followed by units such as 10 μ/mL) across forums on the OHC^2^.

### Generating Responses Using LLMs

We used the OpenAI Chat Completion API to access GPT-3.5-Turbo and GPT-4o. DeepSeek-R1 was accessed through the DeepSeek API and Mistral-7B-Instruct-v0.2 through Hugging Face. We selected these four LLMs to represent a mix of commercial – GPT-3.5-Turbo and GPT-4o – and open-source – DeepSeek-R1 and Mistral-7B-Instruct-v0.2^18–20^ – models that have been commonly evaluated in prior studies^e.g.,5^. After pilot testing multiple prompt formulations (see supplementary materials for details), we observed that small changes in wording and structure produced meaningful differences in LLM responses, which led us to use the original user-posted questions as fixed prompts to ensure consistency and reduce prompt-induced variability across models. This process was intended to assess the different LLMs’ ability to respond to real-world health-related questions about laboratory test results as they are formulated *in situ*. In addition, directly using the original questions preserved the diverse questioning practices and linguistic patterns used by patients and caregivers with varying general and AI literacy levels, while ensuring that both LLM-generated and peer-generated responses were produced in reaction to identical inputs, allowing fair comparison between them.

### Data Analysis

We defined our evaluation metrics based on our research aim and existing literature. Previous research has evaluated LLMs for patient-facing applications as shown in Table 1, including accuracy, currency, consistency with medical standards, clarity, effectiveness, emotional support, transparency, trustworthiness, safety, readability, and personalization. In this analysis, we focus on readability, emotional support, transparency, and personalization, with the primary contributions of our evaluation being a consideration of readability and a more fine-grained view of emotional support. Readability refers to reading difficulty of the text (e.g., use of large words)^21^. It has been less commonly evaluated in studies of LLMs in the medical context, but is a critical metric, as low readability has been associated with low comprehension^21^, which limits use and usefulness of the information. In addition, we take a more granular view of emotional support to understand whether the types of emotional support (e.g., reassurance) offered by humans versus LLMs differ. This could offer insights into the role of humans and LLMs in supporting patients.

**Table 1.** The different ways that patient-facing LLMs have been evaluated in prior research.

| Evaluation Dimensions | Metrics |
| --- | --- |
| Accuracy | numeric scale<sup>22</sup>, qualitative scale<sup>23</sup>, categorical<sup>24</sup> |
| Currency | binary<sup>10</sup> |
| Consistency with medical standards | categorical<sup>23,25</sup>, quantitative metrics<sup>26</sup> |
| Clarity | numeric scale<sup>22,27</sup> |
| Effectiveness | numeric scale<sup>22,27</sup>, qualitative scale<sup>10</sup>, categorical<sup>25</sup> |
| Emotional support | binary<sup>28</sup> |
| Transparency | binary<sup>10,24</sup> |
| Trustworthiness | binary<sup>10</sup>, scale<sup>27</sup> |
| Safety | likelihood of harm and extent of harm<sup>23,25</sup>, quality scale<sup>26</sup>, categorical<sup>24</sup> |
| Readability | computational<sup>10</sup> |
| Personalization | categorical<sup>24</sup> |

#### Computational Analysis of Readability

Following prior work in assessing the readability of medically relevant texts for laypeople^29^, we adopted two validated readability metrics: the Automated Readability Index (ARI), which estimates reading level based on word and sentence length, and the Simple Measure of Gobbledygook (SMOG) score, which estimates reading level based on the frequency of polysyllabic words^30^. We used Carmine DiMascio’s implementation^31^. We applied the readability metrics on both LLM-generated responses and user comments to the original questions. The two metrics can only be calculated when the input texts are at least 100 words, consequently, 52 of the 571 OHC posts being excluded. A one-way ANOVA followed by Tukey’s HSD was applied to evaluate differences in ARI and SMOG readability metrics across five response types (human and four LLMs). All analyses were performed using a significance level of p<0.05.

#### Qualitative Analysis of Emotional Support, Personalization, and Transparency

An initial codebook was created based on the existing literature^**2**^, the evaluation metrics we had decided upon, and an initial review of human and GPT-3.5-Turbo-1106 replies to 122 posts by the research team. Using the codebook, two coders independently coded a subset of 60 OHC user responses (approximately 10% of the sample) across four dimensions: Personalization (e.g., alignment between what is requested and what is provided – low, mid, high), Emotional Support Scale (low, mid, or high), Emotional Support Type (e.g., encouragement, reassurance), and Transparency (e.g., provides references, level of certainty; low, mid, high), for both human and LLM responses. Discrepancies were resolved through iterative discussion with the research team. Once consensus was reached, the two coders recoded the initial subset and then split the remaining 459 human responses and 122 GPT-3.5 Turbo responses (each coding 291 responses).

RD then conducted a quality check by reviewing all original posts, responses, and coding to ensure alignment with the codebook. This same researcher coded all responses from the remaining three LLMs (N = 366), periodically cross-checking coding decisions against the original jointly coded subset to confirm consistency with the initial coding. MAH conducted a final quality review of all coded responses and made the final coding decisions, adjusting around 70 of the 366 items. Final inter-rater agreement was greater than 85% across codes. The final codebook is presented in the results and in supplementary materials.

## Results

LLM-generated responses were significantly longer compared to OHC user replies. User-generated responses averaged 5.09 sentences (SD=4.46). In comparison, GPT-3.5-Turbo-generated responses averaged 9.26 sentences (SD=3.86) (t=-19.06, p<0.005), GPT-4o 15.83 sentences (SD=5.88) (t=-38.21 p<0.005), DeepSeek-R1 18.92 sentences (SD=6.89) (t=-44.52, p<0.005), and Mistral-7B 13.06 sentences (SD=4.67) (t=-32.24, p<0.005).

### Readability

Table 2 summarizes the readability scores for two metrics: ARI and SMOG, across original posts, user comments, and responses generated by four LLMs: GPT-3.5 Turbo, GPT-4o, DeepSeek-R1, and Mistral-7B-Instruct. The readability of the sampled original posts from the OHC showed an ARI of 13.18 (SD=19.09) and a SMOG score of 12.41 (SD=3.77), indicating that these posts required approximately a high school reading level. User comments had similar average scores, with an ARI of 12.09 (SD=6.09) and a SMOG score of 12.54 (SD=2.84), suggesting that peer-generated responses were written in a comparable reading level.

**Table 2.** ARI and SMOG readability scores for original posts, user comments, and LLM-generated responses.

|  |  | ARI Average (STD) | SMOG Average (STD) |
| --- | --- | --- | --- |
| **Original Post** |  | 13.18 (19.09) | 12.41 (3.77) |
| **Human Comment** |  | 12.09 (6.09) | 12.54 (2.84) |
| **LLM- Models** | **GPT-3.5 Turbo** | 13.83 (2.19)<sup>†</sup> | 15.02 (1.50)<sup>*†</sup> |
|  | **GPT-4o** | 13.11 (1.99) | 14.30 (1.33)<sup>*†</sup> |
|  | **DeepSeek-R1** | 12.16 (2.16) | 13.71 (1.49)<sup>*†</sup> |
|  | **Mistral-7B-Instruct** | 12.93 (2.46) | 14.30 (1.65)<sup>*†</sup> |
| ANOVA results: ARI, $F = 14.25$, $p < .001$; SMOG, $F = 17.42$, $p < .001$. | | | |
| *Statistically significant difference compared with the original post based on ANOVA and Tukey's HSD test at $p < 0.05$. | | | |
| †Statistically significant difference compared with human comments based on ANOVA and Tukey's HSD test at $p < 0.05$. | | | |

Mean ARI and SMOG scores corresponded to high school-level reading complexity across models. Both metrics showed a similar relative pattern, with GPT-3.5 Turbo producing the most complex responses with a mean score of 13.83 (SD=2.19), whereas DeepSeek-R1 produced the least complex LLM responses with a mean score of 12.16 (SD=2.16). Based on ARI scores, most LLM responses matched the readability of the original posts and were slightly more complex than human responses.

In contrast, SMOG scores indicated higher complexity for all LLM responses compared with both original posts and human responses. An ANOVA identified significant group differences for both ARI (F=14.25, p<0.001) and SMOG (F=17.42, p<0.001). Post-hoc Tukey tests showed that GPT-3.5 Turbo produced responses requiring a higher reading level than human responses, while GPT-4o, DeepSeek-R1, and Mistral-7B-Instruct did not differ significantly. Among LLMs, GPT-3.5 Turbo differed significantly from DeepSeek-R1 on both metrics, with no other significant pairwise differences. These findings indicate that GPT-3.5 Turbo responses may be less accessible to some patients due to higher reading demands, whereas other models more closely resemble human responses in readability.

### Emotional Support

Table 3 summarizes the qualitative analysis of the level of emotional support provided in human and LLM responses to patient questions related to laboratory test results. Surprisingly, we found that a higher percentage of human responses were rated as providing low levels of emotional support compared to any of the LLMs (Low emotional support: Human=38.4% vs. the highest LLM, GPT-3.5 Turbo=20%). Similarly, human responses were rated to have high levels of emotional support less often compared to LLMs and GPT-3.5 Turbo having the lowest percentage of responses rated as high among the LLMs (High emotional support: Human=49.2% vs. GPT-3.5 Turbo=65.0%). These differences become clear when looking at responses to a post from a user who described severe anxiety and panic after a thyroid medication change, writing, *“I am not functioning well at all and am having terrible anxiety and even panic attacks… Please help*.*”* A peer user replied by sharing personal experience and offering reassurance, saying, *“I know that when you’re hypo… I hope you wake one day feeling slightly improved*.*”* This response helped normalize the experience and reduced the sense of isolation. LLM responses tended to use more direct emotional validation. For instance, Deepseek-R1 began by acknowledging the user’s emotional state: *“It sounds like you’re going through a very challenging time, and I’m sorry to hear about your struggles,”* and then reassured the user by explaining why the symptoms might be temporary. GPT-3.5 Turbo also recognized the user’s distress, stating, *“It sounds like you are going through a tough time… take care of yourself and seek support,”* though its support relied more on general encouragement than explicit emotional support. In reflecting on the styles of the different LLMs, GPT-4o, Deepseek-R1, and Mistral-7b-Instruct were often labeled as providing a ‘High’ level of emotional support and often displayed more direct or strongly supportive language compared to GPT-3.5 Turbo.

**Table 3.** Distribution of Emotional Support Scales in Human-comments vs. LLM models.

| Dimension | Label | Human (N=380) | GPT-3.5 Turbo (N=120) | GPT-4o (N=122) | Deepseek-R1 (N=122) | Mistral-7b-Instruct (N=122) |
| --- | --- | --- | --- | --- | --- | --- |
| Emotional Support Scale | High | 187 (49.2%) | 78 (65.0%) | 87 (71.9%) | 86 (71.1%) | 86 (71.1%) |
|  | Mid | 47 (12.4%) | 19 (15.8%) | 18 (14.9%) | 19 (15.7%) | 18 (14.9%) |
|  | Low | 146 (38.4%) | 24 (20.0%) | 17 (14.0%) | 17 (14.0%) | 18 (14.9%) |
|  | N/A | 139 | 2 | 0 | 0 | 0 |
| *NOTE: The column percentages are based on the number of responses that were applicable. For example, there were 519 human responses to the 122 original posts (519-139=380). |  |  |  |  |  |  |
| *Not Applicable (N/A) indicates the response did not provide emotional support, such as purely informational replies. |  |  |  |  |  |  |

Table 4 summarizes the types of emotional support provided. We identify three key insights from these results: (1) human responses offered the most diverse emotional support, with comfort as the most prevalent type; (2) the other three LLMs primarily provided encouragement, with limited use of other support types; and (3) among LLMs, DeepSeek provided the most diverse emotional support, though encouragement remained its dominant strategy. For example, individuals articulating feelings of distress and despair such as, *“I am too depressed and disturbed by the situation and anxiously waiting*…*”* or *“Every day, I attempt to maintain the same demeanor for my children and fulfill my responsibilities at work. At night, I find myself unable to cease crying, questioning, ‘Where is the fairness in this world?’”*—typically received empathetic responses from peers in the OHC such as, *“I understand the feeling of being overwhelmed*…*”* Peers also provided reassurance through statements such as, *“I do not believe you need to worry,”* and *“You did nothing wrong!”* Comforting language was also utilized to mitigate anxiety: *“Try to remain calm”* and *“Please, take a moment to relax!”*

**Table 4.** Distribution of Emotional Support Types in Human-comments vs. LLM models.

| Dimension | Label | Human (N=341) | GPT-3.5 Turbo (N=122) | GPT-4o (N=121) | Deepseek-R1 (N=121) | Mistral-7b-Instruct (N=122) |
| --- | --- | --- | --- | --- | --- | --- |
| Emotional Support Types | Empathy | 44 (12.9%) | 11 (9%) | 7 (5.8%) | 26 (21.5%) | 6 (4.9%) |
|  | Encouragement | 45 (13.2%) | 90 (73.8%) | 90 (74.3%) | 52 (42.9%) | 100 (82%) |
|  | Comforting | 151 (44.3%) | 9 (7.4%) | 11 (9.1%) | 22 (18.2%) | 9 (7.4%) |
|  | Reassurance | 99 (29%) | 8 (6.5%) | 10 (8.3%) | 18 (14.9%) | 5 (4.1%) |
|  | Sympathy | 2 (0.6%) | 4 (3.3%) | 3 (2.5%) | 3 (2.5%) | 2 (1.6%) |
|  | N/A | 178 | 0 | 1 | 1 | 0 |
| *Not Applicable (N/A) indicates response did not contain an identifiable emotional support type (e.g., informational replies). |  |  |  |  |  |  |

In comparison, while all LLMs typically provided high levels of emotional support, they tended to provide it in a more generic way. Across models, the predominant form of emotional support offered by these responses was encouragement, primarily focusing on proactive health management. For example, an individual posted, *“I am scheduled for a hysterectomy. I recently had a CA 125 the result was 66… I am lost, confused… I can’t even talk about it to my family. All kinds of thoughts are going through my mind, negative thoughts. I don’t know if I should have the surgery or not,”* clearly expressing emotional turmoil and distress. The GPT-3.5 Turbo model responded, *“I understand that you are feeling anxious and overwhelmed… it’s completely normal to feel this way… you are not alone, and there are people and resources available to support you through this journey*.*”*

There were also key differences between the responses generated by different models. To illustrate these differences, we show the LLMs’ responses to a single question from an individual hoping that she is pregnant: *“… Here is what has happened so far: 4 weeks (10/20) HCG = 524, 4 weeks 3 days (10/23) HCG = 1,993, 4 weeks 6 days (10/26) HCG = 7,024. An ultrasound at exactly 5 weeks (10/27) showed a sac but no fetal pole or heartbeat… My questions are: Is all this normal? When can a fetal pole be seen? When can a heartbeat be seen? Are HCG levels normal?”* DeepSeek’s response showed a comforting approach saying *“Congratulations on your pregnancy! It is completely normal to feel anxious during these early stages, especially when waiting for confirmation of a healthy pregnancy…”* By starting with congratulations and normalizing anxiety, it created a supportive atmosphere for the user. In contrast, GPT-4 focused on reassurance and validating the user’s feelings: *“Hi, it’s understandable to have concerns and questions during early pregnancy…”* Mistral-7B answered the same question with encouragement, offering assistance and openness to further questions: *“Hello, I’m here to help answer any questions you have about your pregnancy… I wish you all the best with your pregnancy. If you have any further questions, please don’t hesitate to ask*.*”* These variations in responses reflected distinct approaches to emotional support across models.

### Personalization to Individual Needs

Personalization refers to how much a response addresses an individual’s specific circumstances, needs, and concerns, as expressed in the original post. Interestingly, a lower percentage of human responses were categorized as highly personalized compared to the LLMs (Human = 64% vs. LLM > 90%). Highly personalized human responses typically incorporated specific details regarding the patient’s condition and directly engaged with contextual information from the original post. For instance, one OHC user responded, *“Your husband is not doing himself any favors by continuing to drink…Heavy alcohol consumption with Hepatitis C can expedite disease progression. If he wishes to preserve his liver, he should cease drinking immediately!”* This exemplifies significant engagement with the patient’s unique situation, as the original post described a husband with Hepatitis C who continued drinking alcohol, and the response directly referenced this context while addressing the poster’s primary concern. Conversely, human responses characterized by low personalization tended to offer vague encouragement without directly addressing the poster’s specific concerns. For example, in response to a detailed and emotionally vulnerable post describing worsening anxiety, fatigue, and thyroid fluctuations following about of stomach flu, one reply simply stated, *“I know what you are going through. I suffer from severe dizziness*…*I wish you the best*.*”*

In contrast, LLM-generated responses were consistently categorized as demonstrating high personalization, like providing emotional support, the personalization had a distinct pattern. All the LLM responses acted as a mirror, reflecting the user’s specific symptoms or test results back to them in a coherent and organized manner. This pattern was largely consistent across models, with only minor differences in the amount of detail included. For example, in response to a post from a male (age 41-45), seeking expert advice on his thyroid diagnosis and treatment, GPT-3.5 Turbo crafted a response that began by empathetically acknowledging the user’s specific concern: *“It is understandable that you harbor concerns regarding your thyroid levels and the symptoms you are experiencing*…*”* The user had shared a detailed medical history, noting a persistently high TSH level (rising from 7.52 to 47.15 within a year), normal T3 and T4 values, and symptoms such as fatigue, poor concentration, palpitations, and lack of interest. He also mentioned confusion about the increase in his thyroxine medication dosage from 25mcg to 100mcg without much explanation from his physician and expressed concern about what might be happening to his metabolic system. In response, LLMs (all four) reflected back the user’s concern and offered tailored advice and health recommendations. For example, GPT-3.5 Turbo stated, *“*… *Lifestyle adjustments, including the maintenance of a balanced diet, stress reduction, and regular physical activity, may also contribute to the enhancement of your thyroid health*…*”*

However, among high personalization LLM responses, there were clear differences in how models personalized their replies. DeepSeek tended to personalize responses through detailed, condition-specific explanations that closely reflected the user’s reported test results. In contrast, ChatGPT-4.0 relied on broader explanatory framing, situating the user’s concern within general medical information rather than focusing narrowly on specific values. Mistral-7B adopted a more constrained personalization approach, acknowledging the user’s situation while clarifying role boundaries before offering general guidance. These patterns correspond to the distribution of personalization observed across models, as summarized in Table 5. Across models, personalization was achieved through different mechanisms, including close reflection of reported symptoms, condition-specific explanation, and clarification of informational scope, even when responses relied primarily on details provided in the original post.

**Table 5.** Distribution of Personalization in Human-comments vs. LLM models.

| Dimension | Label | Human Comments (N=383) | GPT-3.5 Turbo (N=121) | GPT-4o (N=122) | Deepseek-R1 (N=122) | Mistral-7b-Instruct (N=122) |
| --- | --- | --- | --- | --- | --- | --- |
| Personalization | High | 245 (64.0%) | 118 (97.5%) | 111 (91.0%) | 117 (96.0%) | 116 (95.1%) |
|  | Mid | 46 (12.0%) | 1 (0.8%) | 1 (0.8%) | 1 (0.8%) | 1 (0.8%) |
|  | Low | 92 (24.0%) | 2 (1.7%) | 10 (8.2%) | 4 (3.2%) | 5 (4.1%) |
|  | N/A* | 136 | 1 | 0 | 0 | 0 |
| *Not Applicable (N/A) indicates personal experiences that were not answering the original poster’s question (e.g., piggybacking with their own question). |  |  |  |  |  |  |

Finally, not all LLM responses demonstrated a high level of personalization. In some cases, the personalization was low and limited to restating the information provided by the user without offering deeper context, tailored advice, or emotional support. For example, one response to a user who posted about their Hepatitis C viral load simply reiterated the lab result and offered a generic recommendation: *“…your HCV RNA RT-PCR test has identified a viral load of 324,000 IU/mL… It is vital to discuss these results along with your overall diagnosis with your healthcare provider…”*

### Transparency

When we evaluated transparency in responses, we considered whether the information is clear, logically structured, and supported by meaningful explanations of reasoning. Overall, human responses were more likely to be rated as highly transparent compared to any of the LLMs (Human = 51.6% vs. LLMs < 33%; Table 6). Even so, the human responses varied widely in their transparency. Highly transparent human responses often explained the reasoning behind medical interpretations step by step, clarified why specific tests were emphasized, and linked conclusions to treatment decisions. For example, one user explained why their clinician prioritized Free T3 and Free T4 over TSH, noting that *“… from what I’ve read, TSH changes often with Hashi. My endo doesn’t place as much value on the TSH as he does on the FT3/FT4 levels because the frees show directly what the thyroid is doing, and the TSH is a pituitary hormone. He mainly treats the frees and my symptoms…”* The response also directed the user to a vetted resource for specialized care, providing both the recommendation and its rationale.

**Table 6.** Distribution of Transparency in Human-comments vs. LLM models.

| Dimension | Label | Human Comments (N=343) | GPT-3.5 Turbo (N=117) | GPT-4o (N=122) | Deepseek-R1 (N=122) | Mistral-7b-Instruct (N=122) |
| --- | --- | --- | --- | --- | --- | --- |
| Transparency | High | 177 (51.6%) | 38 (32.5%) | 35 (28.7%) | 37 (30.0%) | 39 (32.0%) |
|  | Mid | 10 (2.9%) | 0 | 0 | 0 | 0 |
|  | Low | 156 (45.5%) | 79 (67.5%) | 86 (70.5%) | 85 (70.0%) | 83 (68.0%) |
|  | N/A* | 176 | 5 | 0 | 0 | 0 |
| *Not Applicable (N/A) responses focused less on providing medical information and more on offering emotional support, sharing personal experiences, or engaging in normal conversation. |  |  |  |  |  |  |

Other human responses relied on personal opinions, observations, and experiences and cite vague sources or do not offer supporting evidence at all. For instance, one OHC user stated: *“From what I have heard and read, PegIntron is a better choice for tough strains of Geno 1—although doctors will always have their own opinions, and that is just how they usually decide. WHICH treatment you go on is the one that their center supports the drug company of. I have Geno 1A and also 1B, and two doctors insisted on Intron…”* While this individual refers to sources, they are not specific and do not provide links for the original poster to learn more and draw their own conclusions.

The four LLMs were rated similarly for their transparency, with GPT-3.5 Turbo generating the highest percentage of high transparency responses and GPT-4o the lowest (32.5% vs. 28.7%). LLM-generated responses again exhibited distinct patterns compared to peer patient comments, particularly in how they structured and conveyed information. While the LLM responses were often detailed and well-organized, and transparent in their limits as an AI tool, they frequently lacked supporting evidence, in-depth explanations of reasoning, used cautious language, and general statements aimed at neutrality. This sometimes resulted in vague or less useful responses. For instance, GPT-3.5 Turbo acknowledged the complexity of the patient’s symptoms but remained nonspecific and lacked concrete guidance: *“It sounds like you have been experiencing a wide range of symptoms… it’s important to consider further investigation into your thyroid health… it’s important to advocate for yourself and seek the care and attention you need*.*”* High transparency LLM-generated responses provided clear explanations and step-by-step reasoning.

## Discussion

Previous studies have demonstrated that LLMs provide medically accurate and safe responses to patient questions and perform well in clinical reasoning and evidence summarization tasks^5,6,9^. Our study extends this literature by systematically evaluating patient-centered dimensions, including personalization, emotional support, transparency, and readability, across multiple LLMs and contrasting them with human responses. Together, these dimensions provide insight into the potential role of LLMs within the broader ecosystem of support in OHCs. Below, we situate our findings in relation to prior work and discuss implications for LLM developers and OHC platforms.

### Comparison with Previous Literature

While previous studies have shown the potential of LLMs in providing emotional support^28^, a key finding from our study is a more nuanced view of this support. Peer users provided diverse forms of emotional support, including comfort, reassurance, and empathy, whereas LLM responses tended to rely on more generic expressions, most often encouragement. Prior research in health communication and patient support consistently emphasizes the importance of empathy and emotional validation in reducing anxiety, fostering trust, and supporting coping during illness and treatment^14,28^. Human empathy has been shown to improve patient satisfaction, perceived support, and engagement, particularly in contexts involving uncertainty or chronic conditions^28^. Our findings suggest that current LLMs do not yet replicate these relational aspects of peer support.

Enhancing emotional responsiveness in LLMs therefore represents an important opportunity for improving their usefulness in patient support contexts. Prior work in conversational agents and mental health chatbots suggests that models trained on emotionally rich data and guided by explicit affective objectives can improve perceived empathy and user satisfaction^23,24^. Techniques such as fine-tuning on peer support interactions, incorporating emotion recognition, and designing prompts or system instructions that encourage acknowledgment of emotional cues may help LLMs respond more sensitively to individual emotional needs^28^. While further evaluation is needed, this literature suggests feasible pathways for improving emotional alignment.

Beyond emotional support, prior literature identifies personalization as a key dimension of effective AI-based health support. Studies show that personalized systems improve relevance and engagement by adapting content to users’ health conditions, goals, and prior interactions^14^. Health chatbots typically rely on explicit user input to tailor information, most often through content adaptation rather than interface changes^23^. More advanced systems integrate personal health data, lifestyle information, and interaction history to support longitudinal engagement and deliver more context-aware guidance^13^. This aligns with our findings, where responses referenced users’ individual context and prior disclosures, whereas generic responses did not. Together, these studies position personalization as an important complement to emotional support in shaping user experience and trust in AI-mediated health interactions.

Transparency has also been emphasized as a critical requirement for AI-mediated health communication. Prior research shows that patients expect clear disclosure of AI involvement, understandable explanations of how responses are generated, and explicit communication of system limitations and human oversight^28^. Opacity in AI systems has been linked to reduced trust and weaker informed decision-making, particularly in clinical contexts where users rely on AI output to interpret health information^23^. Interactive conversational designs that allow users to ask follow-up questions further strengthen understanding and trust^25^. These findings align with our results by reinforcing the importance of visible reasoning and accountability in patient-facing LLM systems.

### Role of LLMs in Online Health Communities

OHCs face ongoing challenges related to variability in response quality, delayed replies, and uneven access to medically credible information^16^. In this context, LLMs may be particularly useful for providing timely explanations of laboratory test results, summarizing medical concepts in accessible language, or offering initial guidance when peer responses are unavailable or incomplete^28^. Our findings suggest that LLMs may be especially valuable when informational needs are unmet by peers, while human responders remain critical for emotionally nuanced support.

Personalization emerged as a key point of difference between LLM and human responses. Human responders often personalized advice through lived experience and prior knowledge. LLMs personalized responses by restating and organizing details from the original post. This approach produced consistently high personalization scores for LLMs, but it lacked the experiential grounding seen in many peer replies. Human responses frequently incorporated personal histories, firsthand insights, and contextual nuance that went beyond the information explicitly provided. Expanding LLM access to longitudinal context, including prior posts, historical laboratory results, and patient-stated preferences, offers a clear path to stronger personalization while preserving informational accuracy^23,28^.

Building on these findings, an adaptive support model could combine the complementary strengths of LLMs and peer responders. Minimum standards for response timeliness and support coverage could guide system behavior. For example, when a post receives no response within a defined time window, an LLM could provide an initial informational reply. When peer responses focus primarily on emotional support, an LLM could supplement them with factual clarification or structured guidance. When only an LLM response is present, the system could prompt peer engagement by highlighting the post to relevant community members or recommending similar peer experiences. Such an adaptive approach operationalizes the roles identified in our results by aligning emotional support, personalization, and transparency with the timing and type of responses delivered.

For example, in some HIV-related discussions, we observed that peer users frequently offered emotional reassurance but often lacked detailed medical information. In such situations, LLMs could complement peer responses by supplying accurate, evidence-based explanations while peers continue to provide relational and emotional support. This division of labor highlights the potential for LLMs to augment, rather than replace, human participation in OHCs. At the same time, privacy considerations shape how and when such augmentation is appropriate. Prior work on OHCs shows that patients often choose peer spaces specifically to share sensitive experiences in environments perceived as human, empathetic, and socially safe^23,28^. Motivations for engaging with peers include seeking lived experience, social validation, and a sense of belonging, particularly for stigmatized conditions such as HIV. These needs are closely tied to trust and control over personal disclosures, which automated systems may not fully replicate. This reinforces the importance of limiting LLM involvement to clearly defined supportive roles, while preserving peer spaces for experiential exchange and identity-based support. Rather than replacing peer interaction, LLMs should operate in ways that respect privacy expectations and maintain the social functions that motivate participation in OHCs.

### Integrating LLMs into Online Health Communities for Holistic Patient Support

As mentioned, LLMs should be viewed as a part of a patient’s support ecosystem and the goal should be to enhance peer engagement and support, rather than supplant it. Optimizing readability, improving contextual personalization, and refining emotional responsiveness may allow LLMs to play a meaningful role in patient support on OHCs. From a development perspective, OHC data could inform fine tuning strategies to better reflect real patient concerns, while retrieval augmented generation using biomedical literature may reduce hallucinations and improve transparency. Thoughtful integration of these approaches may help align LLM capabilities with the complex informational and emotional needs present in online health communities^23,28^.

Building on these, LLMs should be designed to intervene selectively when gaps in peer support emerge, such as delayed responses or unmet informational needs^23^. Designers should prioritize task-specific functions including explanation, summarization, and clarification, while relying on peers for experiential and relational support^9,23,28^. Personalization should emphasize contextual accuracy through longitudinal user data rather than simulated lived experience. Emotional responsiveness should focus on acknowledging distress and guiding users toward peer engagement. Finally, transparent disclosure of AI involvement, clear communication of limitations, and alignment with community governance norms are essential for sustaining trust and appropriate use in OHC settings.

Based on prior work and our findings, LLMs do not need to replicate all functions of human peers to be suitable for integration into OHCs, but they must meet minimum standards aligned with their intended role^28^. At a minimum, LLMs should provide accurate, clearly sourced information; disclose their non-human status and limitations; respond in a timely manner when informational gaps exist; and respect community norms through constrained, context-aware participation. Achieving this requires models that are trained or adapted for health contexts rather than relying solely on general-purpose LLMs^25^. Practical steps include fine-tuning on OHC-relevant data, integrating retrieval from vetted biomedical sources, enabling access to longitudinal context, and embedding governance rules that determine when and how the model intervenes. These requirements position LLMs as reliable, bounded contributors rather than replacements for peer support.

Integrating LLMs and AI into OHCs should also respect established community norms and governance. Some communities explicitly prohibit AI generated content due to risks of misinformation and potential harm. For example, the r/diabetes community states, *“AI tools are incapable of vetting the information they put out as being accurate and factual… Because the information AI generates has the potential to be wildly false, we do NOT allow posts or comments to utilize AI output as general advice or presenting it as information*.*”*^32^ These positions highlight the importance of accounting for user and moderator attitudes, maintaining transparency about AI involvement, and implementing safeguards to prevent misuse. Effective integration will require platforms to label AI-generated and AI-assisted replies, make AI assistance opt-in, and let each community set its own disclosure norms rather than impose them platform-wide.

### Limitations and Future Work

This study has several limitations that should be considered when interpreting the findings. First, we evaluated a limited set of LLMs rather than the full range of available generative models. As a result, our findings may not generalize to other models with different architectures, training data, or alignment strategies. We evaluated only general-purpose LLMs and no medically fine-tuned or domain-specific models. This reflects our focus on the widely available tools patients’ access, since specialized systems remain out of reach for most laypeople due to cost or lack of awareness. Domain-specific models may perform differently and benchmarking them against general-purpose tools is an important direction for future work. Second, we used original OHC posts as prompts without additional prompt engineering and restricted the analysis to single interactions. This approach was chosen to mirror how patients or caregivers might naturally seek assistance when encountering unfamiliar laboratory results. However, it does not capture how LLM performance might change with iterative clarification, follow up questions, or optimized prompting. Limiting the analysis to single interactions may therefore underestimate the potential capabilities of LLMs in more interactive settings. This should be explored in future *in situ* work with OHC users. Third, similar to past research, this study focused on data from a single OHC^2^. This limits the generalizability of the findings to other platforms, conditions, or patient populations. Replication across multiple OHCs will be important for assessing robustness and transferability.

## Conclusion

Human and LLM responses to questions about laboratory test results show clear and complementary strengths. Peer responses offer rich emotional support and lived experience, though they vary in reliability, transparency, and the level of detail provided. LLMs, in contrast, consistently produce clear, structured, and contextually organized information, but they remain limited in emotional nuance, transparency of reasoning, and sensitivity to community norms. For these reasons, general-purpose LLMs that lack domain adaptation and governance safeguards are not yet ready for direct integration into online health communities. As these systems improve, particularly in contextual awareness, transparency, and emotional responsiveness, they hold promise as supportive tools that expand access to reliable information while working alongside, rather than replacing, peer-based support in OHCs.

## Data Availability

Not Available

## Acknowledgements

We are grateful to Zipporah Jones-Hamlin and Tartela Tabassum for their early contributions to the coding.

## References

1. Caldeira C, Gui X, Reynolds TL, Bietz M, Chen Y. Managing healthcare conflicts when living with multiple chronic conditions. International Journal of Human-Computer Studies. 2021;145:102494.

2. Reynolds TL, Zhang J, Zheng K, Chen Y. Unpacking the Use of Laboratory Test Results in an Online Health Community throughout the Medical Care Trajectory. Proc ACM Hum-Comput Interact. 2022;6(CSCW2):1–32.

3. Young AL, Miller AD. “This Girl is on Fire”: Sensemaking in an Online Health Community for Vulvodynia. In: Proceedings of the 2019 CHI Conference on Human Factors in Computing Systems. ACM; 2019:1–13.

4. Holohan M. A boy saw 17 doctors over 3 years for chronic pain. ChatGPT found the diagnosis. TODAY.com. September 12, 2023. https://www.today.com/health/mom-chatgpt-diagnosis-pain-rcna101843 [accessed 5/9/24]

5. He Z, Bhasuran B, Jin Q, et al. Quality of Answers of Generative Large Language Models Versus Peer Users for Interpreting Laboratory Test Results for Lay Patients: Evaluation Study. J Med Internet Res. 2024;26:e56655. doi:10.2196/56655

6. Chen S, Guevara M, Moningi S, et al. The effect of using a large language model to respond to patient messages. The Lancet Digital Health. 2024;6(6):e379–e381. doi:10.1016/S2589-7500(24)00060-8

7. Aljamaan F, Temsah MH, Altamimi I, et al. Reference Hallucination Score for Medical Artificial Intelligence Chatbots: Development and Usability Study. JMIR Med Inform. 2024;12:e54345. doi:10.2196/54345

8. Mugaanyi J, Cai L, Cheng S, Lu C, Huang J. Evaluation of Large Language Model Performance and Reliability for Citations and References in Scholarly Writing: Cross-Disciplinary Study. J Med Internet Res. 2024;26:e52935. doi:10.2196/52935

9. Tang L, Sun Z, Idnay B, et al. Evaluating large language models on medical evidence summarization. npj Digit Med. 2023;6(1):158. doi:10.1038/s41746-023-00896-7

10. Hristidis V, Ruggiano N, Brown EL, Ganta SRR, Stewart S. ChatGPT vs Google for Queries Related to Dementia and Other Cognitive Decline: Comparison of Results. J Med Internet Res. 2023;25:e48966.

11. Morris TA, Guard JR, Marine SA, et al. Approaching Equity in Consumer Health Information Delivery: Net Wellness. JAMIA. 1997;4(1):6–13. doi:10.1136/jamia.1997.0040006

12. Reynolds TL, Ali N, McGregor E, et al.Understanding Patient Questions about their Medical Records in an Online Health Forum: Opportunity for Patient Portal Design.

13. Zhang Zhan, Lu Yu, Kou Yubo, Wu Danny T.Y., Huh-Yoo Jina, He Zhe. Understanding Patient Information Needs About Their Clinical Laboratory Results: A Study of Social Q&A Site. In: Studies in Health Technology and Informatics. IOS Press; 2019. doi:10.3233/SHTI190458

14. Chuang KY, Yang CC. Helping you to help me: Exploring supportive interaction in online health community. Proc of Assoc for Info. 2010;47(1):1–10. doi:10.1002/meet.14504701140

15. Giardina TD, Baldwin J, Nystrom DT, Sittig DF, Singh H. Patient perceptions of receiving test results via online portals: a mixed-methods study. Journal of the American Medical Informatics Association. 2018;25(4):440–446. doi:10.1093/jamia/ocx140

16. Sun S, Zhou X, Denny JC, Rosenbloom T, Xu H. Understanding patient-provider communication entered via a patient portal system. Proc of Assoc for Info. 2012;49(1):1–4. doi:10.1002/meet.14504901387

17. Zikmund-Fisher BJ, Exe NL, Witteman HO. Numeracy and Literacy Independently Predict Patients’ Ability to Identify Out-of-Range Test Results. J Med Internet Res. 2014;16(8):e187. doi:10.2196/jmir.3241

18. Luo D, Liu M, Yu R, et al. Evaluating the performance of GPT-3.5, GPT-4, and GPT-4o in the Chinese National Medical Licensing Examination. Sci Rep. 2025;15(1):14119. doi:10.1038/s41598-025-98949-2

19. Jiang AQ, Sablayrolles A, Mensch A, et al. Mistral 7B. arXiv. Preprint online 2023:arXiv:2310.06825.

20. Sandmann S, Hegselmann S, Fujarski M, et al. Benchmark evaluation of DeepSeek large language models in clinical decision-making. Nat Med. 2025;31(8):2546–2549. doi:10.1038/s41591-025-03727-2

21. Garner M, Ning Z, Francis J. A framework for the evaluation of patient information leaflets. Health Expectations. 2012;15(3):283–294. doi:10.1111/j.1369-7625.2011.00665.x

22. Lahat A, Shachar E, Avidan B, Glicksberg B, Klang E. Evaluating the Utility of a Large Language Model in Answering Common Patients’ Gastrointestinal Health-Related Questions: Are We There Yet? Diagnostics. 2023;13(11):11. doi:10.3390/diagnostics13111950

23. Bernstein IA, Zhang Y (Victor), Govil D, et al. Comparison of Ophthalmologist and Large Language Model Chatbot Responses to Online Patient Eye Care Questions. JAMA Netw Open. 2023;6(8):e2330320. doi:10.1001/jamanetworkopen.2023.30320

24. Kim Y, Lee J, Kim S, Park J, Kim J. Understanding Users’ Dissatisfaction with ChatGPT Responses: Types, Resolving Tactics, and the Effect of Knowledge Level. In: Proceedings of the 29th International Conference on Intelligent User Interfaces. ACM; 2024:385–404. doi:10.1145/3640543.3645148

25. Singhal K, Azizi S, Tu T, et al. Large language models encode clinical knowledge. Nature. 2023;620(7972):172–180. doi:10.1038/s41586-023-06291-2

26. Coskun B, Ocakoglu G, Yetemen M, Kaygisiz O. Can ChatGPT, an Artificial Intelligence Language Model, Provide Accurate and High-quality Patient Information on Prostate Cancer? Urology. 2023;180:35–58.

27. Yang Z, Xu X, Yao B, et al. Talk2Care: Facilitating Asynchronous Patient-Provider Communication with Large-Language-Model. arXiv. Preprint posted online February 3, 2024:arXiv:2309.09357.

28. Saha K, Jain Y, Liu C, Kaliappan S, Karkar R. AI vs. Humans for Online Support: Comparing the Language of Responses from LLMs and Online Communities of Alzheimer’s Disease. ACM Trans Comput Healthcare. Published online January 3, 2025:3709366. doi:10.1145/3709366

29. Wu DT, Hanauer DA, Mei Q, et al. Assessing the readability of ClinicalTrials.gov. Journal of the American Medical Informatics Association. 2016;23(2):269–275. doi:10.1093/jamia/ocv062

30. Mac O, Ayre J, Bell K, McCaffery K, Muscat DM. Comparison of Readability Scores for Written Health Information Across Formulas Using Automated vs Manual Measures. JAMA Netw Open. 2022;5(12):e2246051.

31. DiMascio C. cdimascio/py-readability-metrics. Published online June 17, 2024. Accessed June 20, 2024. https://github.com/cdimascio/py-readability-metrics

32. Diabetes and AI : r/diabetes. Reddit. Accessed January 21, 2026. https://www.reddit.com/r/diabetes/comments/1pdo7jt/diabetes_and_ai

